# Compatibility of National Food Composition Databases with USDA FoodData Central: A Seven-Country LLM-Based Analysis

**DOI:** 10.64898/2026.05.23.26353942

**Authors:** Shin-ichi Nakagawa, Akira Yamamoto

## Abstract

To evaluate the international interoperability of food composition databases, we assessed the compatibility of seven national food composition tables with USDA FoodData Central (FDC) using the LLM-based matching method reported previously (Nakagawa and Yamamoto, 2026). Databases from four English-speaking countries (Canada, United Kingdom, Australia, and New Zealand), South Korea, and Japan were compared with 8,158 USDA FDC entries (SR Legacy and Foundation Foods, excluding Survey/FNDDS). Match rates varied by country (62.0–89.7%) and food category. After excluding six USDA categories unsuitable for cross-national comparison, 45.2% of the remaining 6,290 entries were not matched by any country. Canada showed the highest concordance, reflecting shared North American food supply. Japan and South Korea showed similar low coverage for vegetables and spices. These findings suggest that while USDA FDC represents a practical foundation for a globally comprehensive food composition database given its breadth, systematic incorporation of country-specific foods and classification schemes will be necessary to achieve true international interoperability.

## 1. Introduction

Food composition databases (FCDs) are essential tools in nutritional epidemiology, dietary assessment, and public health research (Greenfield and Southgate, 2003). Cross-national dietary comparisons, however, face a fundamental methodological challenge: nutrient values for equivalent foods frequently differ across national FCDs due to differences in analytical methods, food sampling strategies, cultivar composition, and food processing practices (Sichert-Hellert et al., 2011; Jenab et al., 2009). The most rigorous solution—direct laboratory measurement of all food items in each country—is clearly impractical at scale. As a result, researchers conducting multi-country studies have typically adopted one of two approaches: using a single reference database such as USDA FDC as a surrogate standard (Dehghan et al., 2006; Viera Aller et al., 2015), or developing harmonized multi-country databases through labor-intensive manual food matching (FAO/INFOODS, 2012; Slimani et al., 1999). USDA FoodData Central (FDC) (USDA, 2024) is widely used as a de facto international reference owing to its comprehensiveness and open access. International efforts to harmonize food composition data, notably through FAO/INFOODS and the European Food Information Resource (EuroFIR) project (2005–2013), have established guidelines for food matching and data quality (FAO/INFOODS, 2012), but item-level correspondence between national FCDs remains largely unestablished. In a preceding study, we developed a large language model (LLM)-based matching methodology between the Japanese Standard Tables of Food Composition 2020 and USDA FDC, demonstrating that 22.2% of Japanese food items lacked a USDA equivalent (Nakagawa and Yamamoto, 2026). Here, we extended this methodology to six additional national FCDs to systematically quantify USDA FDC’s international coverage and characterize its gaps across diverse dietary cultures.

## 2. Materials and Methods

### 2.1 Databases

Seven national FCDs were selected based on open data availability and geographic diversity (Table 1): Japan (MEXT, 2020), United Kingdom (PHE, 2021), Australia (FSANZ, 2024), New Zealand (Plant & Food Research, 2024), Canada (Health Canada, 2015), and South Korea (Ministry of Food and Drug Safety, 2026). The USDA FDC (USDA, 2024) reference set comprised 8,158 entries (SR Legacy: 7,793; Foundation Foods: 365), excluding Survey/FNDDS items (n=5,432).

**Table 1.** Matching results for seven national food composition databases against USDA FoodData Central.

| Country | n | Matched, n (%) | No-match, n (%) | High conf. (%) | Mean dE (%) | Database |
| --- | --- | --- | --- | --- | --- | --- |
| JP | 2,478 | 1,539 (62.1) | 939 (37.9) | 78.7 | 4.3 | MEXT 8th ed. (2020) |
| UK | 2,886 | 1,789 (62.0) | 1,097 (38.0) | 73.8 | 7.0 | CoFID (2021) |
| AU | 3,741 | 2,925 (78.2) | 814 (21.8) | 72.6 | 9.3 | AUSNUT 2023 |
| NZ | 2,763 | 1,831 (66.3) | 931 (33.7) | 84.3 | 6.1 | NZFCD (2024) |
| CA | 5,690 | 5,102 (89.7) | 587 (10.3) | 91.8 | 4.0 | CNF 2015 |
| KR | 3,310 | 2,195 (66.3) | 1,114 (33.7) | 83.6 | 4.6 | KFCT DB 10.2 (2026) |

dE: mean absolute percentage difference in energy between matched pairs (items $\geq 5$ kcal/100 g only).
MEXT: Ministry of Education, Culture, Sports, Science and Technology; CoFID: Composition of Foods Integrated Dataset; AUSNUT: Australian Food and Nutrient Database; NZFCD: New Zealand Food Composition Database; CNF: Canadian Nutrient File; KFCT: Korean Food Composition Table.

### 2.2 Nutrient variables

Seventeen nutrient variables common to USDA FDC and all seven national FCDs were used: energy (kcal), protein (g), fat (g), carbohydrate (g), dietary fiber (g), calcium (mg), iron (mg), potassium (mg), zinc (mg), vitamin C (mg), vitamin A as RAE (μg), vitamin B1 (mg), vitamin B2 (mg), vitamin D (μg), vitamin B12 (μg), and salt equivalent (g). These variables were selected as the largest common subset across all databases. Some nutrients showed substantial missing rates in certain databases—notably vitamin D and vitamin B12 in the Korean KFCT (39.6% and 35.3%, respectively) and vitamin D in the Japanese MEXT (3.9%)—but missing values were retained as null rather than excluded, to preserve the full item count for matching.

### 2.3 Matching pipeline

The matching pipeline followed the approach described previously (Nakagawa and Yamamoto, 2026), with country-specific prompt customizations as described below. Briefly: (1) food category pre-filtering using a manually constructed mapping table between each national FCD’s category system and USDA FDC categories; (2) Euclidean distance ranking over the 17 normalized nutrient variables, selecting the top 25 candidates; and (3) LLM-based judgment using Claude Haiku (claude-haiku-4-5-20251001; Anthropic, 2024).

### 2.4 Prompt design

The LLM was queried with a structured prompt for each food item presenting: (a) the source food name and category, (b) its 17-nutrient profile per 100 g, and (c) the 25 USDA candidate entries with their nutrient profiles. The model was instructed to select the single best conceptual and nutritional match, or to designate NO_MATCH if no adequate equivalent existed. Country-specific instructions were added to improve precision; for example, for Japanese foods: “Choose NO_MATCH for Japan-specific foods with no US equivalent (e.g. natto, miso, kombu, wakame, umeboshi, tsukudani, etc.)”. Responses were constrained to JSON format:

{“match_number”: <1–25 or NO_MATCH>, “confidence”: “<high|medium|low>“, “reason”: “<one line>“}

Confidence ratings reflected the LLM’s self-assessed certainty: high indicated a conceptually and nutritionally close match; medium indicated a reasonable but imperfect match; low indicated a tentative match. The complete prompt templates for all seven countries will be made publicly available at https://github.com/shnkgw-rincom/jbfd-correspondence-table upon finalization.

### 2.5 USDA category exclusions

Six USDA categories were excluded from the international coverage analysis a priori due to limited cross-national applicability: American Indian/Alaska Native Foods (n=165, 100% unmatched), Restaurant Foods (n=113, 100%), Meals, Entrees, and Side Dishes (n=81, 100%), Baby Foods (n=345, 75.4%), Beef Products (n=969, 61.0%; excessive US-specific subcategorization), and Breakfast Cereals (n=195, 57.4%; brand-dominated). The remaining analytical dataset comprised 6,290 USDA entries.

## 3. Results

Match rates ranged from 62.0% (UK) to 89.7% (CA), with mean energy differences between matched pairs (dE) of 4.0–9.3% (Table 1). Of 8,158 USDA entries, 3,985 (48.8%) were matched by none of the seven countries. After excluding the six a priori categories (n=1,868), 2,842 of the remaining 6,290 entries (45.2%) were unmatched. No USDA entry was matched by all seven countries simultaneously; 83 entries (1.3%) were matched by six countries and 190 (3.0%) by five countries. Coverage varied systematically by food category (Table 2). Poultry products (mean 95.3%), pork products (92.5%), and sweets (86.8%) showed consistently high match rates. In contrast, spices and herbs (mean 55.2%), fast foods (64.5%), and vegetables (60.7%) showed the lowest and most variable coverage. Canada showed the highest match rates across most categories. The United Kingdom showed notably low rates for fast foods (20.2%) and soups/sauces (27.4%), suggesting that the UK food supply diverges substantially from the US in prepared and composite dishes. Japan and South Korea showed similar low coverage for vegetables (37.6% and 37.7%) and spices/herbs (28.4% and 40.2%), suggesting shared East Asian dietary characteristics. Among the four English-speaking countries, match rates varied considerably across categories, suggesting that shared language does not necessarily imply dietary similarity with the US food supply.

**Table 2.** Match rates (%) by USDA food category for each of the seven countries. “—” indicates that the food category was not represented in the national database classification.

| **USDA Category** | **JP** | **UK** | **AU** | **NZ** | **CA** | **KR** | **Mean** |
| --- | --- | --- | --- | --- | --- | --- | --- |
| Cereal Grains and Pasta | 70.7% | 77.6% | 78.5% | 70.9% | 92.3% | 79.2% | 78.2% |
| Baked Products | — | 74.4% | 96.2% | 96.7% | 96.4% | — | 90.9% |
| Vegetables and Vegetable Products | 37.6% | 52.2% | 70.0% | 63.8% | 84.7% | 37.7% | 57.7% |
| Fruits and Fruit Juices | 69.4% | 56.0% | 65.7% | 68.6% | 91.5% | 73.4% | 70.8% |
| Legumes and Legume Products | 62.0% | 56.1% | 68.7% | 56.8% | 94.3% | 83.3% | 70.2% |
| Nut and Seed Products | 82.6% | 56.0% | 74.3% | 69.5% | 97.8% | 79.1% | 76.5% |
| Beef Products | 84.2% | 81.0% | 81.2% | 86.1% | 94.7% | 88.9% | 86.0% |
| Pork Products | — | 85.7% | 96.7% | 90.0% | 97.6% | — | 92.5% |
| Poultry Products | — | 92.6% | 97.5% | 94.9% | 96.2% | — | 95.3% |
| Lamb, Veal, and Game Products | — | 96.8% | 88.3% | 97.6% | 79.1% | — | 90.4% |
| Sausages and Luncheon Meats | — | 66.7% | 89.4% | 95.2% | 97.3% | — | 87.2% |
| Finfish and Shellfish Products | 74.4% | 71.6% | 91.8% | 69.2% | 84.9% | 70.2% | 77.0% |
| Dairy and Egg Products | 85.4% | 75.7% | 91.4% | 82.1% | 90.0% | 91.3% | 86.0% |
| Fats and Oils | 94.1% | 67.5% | 79.2% | 77.3% | 84.0% | 50.0% | 75.4% |
| Spices and Herbs | 28.4% | 39.3% | 52.9% | 60.7% | 71.4% | 40.2% | 48.8% |
| Soups, Sauces, and Gravies | — | 27.4% | 74.1% | 78.2% | 92.5% | — | 68.0% |
| Sweets | 63.7% | 82.2% | 94.4% | 100.0% | 93.8% | 86.5% | 86.8% |
| Snacks | — | 66.7% | 89.5% | 93.5% | 98.9% | — | 87.2% |
| Beverages | 63.9% | 77.3% | 79.4% | 79.6% | 92.2% | 84.6% | 79.5% |
| Fast Foods | 50.0% | 20.2% | 75.0% | 67.2% | 65.0% | 69.2% | 57.8% |

Mean: average match rate across countries with available data for that category.

## 4. Discussion

This study provides a systematic quantification of USDA FDC’s international coverage across seven national FCDs using a uniform LLM-based methodology. The finding that 45.2% of analytically eligible USDA entries were matched by no country suggests that a substantial portion of USDA FDC reflects US-specific foods not present in other national food supplies.

Previous multi-country dietary studies have addressed the problem of database incompatibility by using USDA FDC as a common reference, supplemented with local food composition data (Dehghan et al., 2006; Viera Aller et al., 2015). While pragmatically effective, this approach implicitly assumes that USDA FDC provides adequate coverage of non-US foods—an assumption our findings call into question. Comprehensive direct measurement of all foods in each country would represent the gold standard for cross-national comparisons, but is clearly impractical given the scale of modern food supplies. LLM-based matching offers a scalable middle ground, enabling systematic identification of coverage gaps without requiring laboratory analysis.

The consistently low match rates for vegetables, spices, and fast foods reflect the cultural specificity of culinary traditions. Japan and South Korea share similarly low coverage in these categories, consistent with the prominence of fermented vegetables, wild plants, and country-specific condiments absent from the US food supply. Conversely, the high and consistent match rates for poultry and dairy products across all countries underscore the universality of these protein sources in global diets. Canada’s markedly high overall match rate (89.7%) reflects shared North American food supply chains and the historical alignment of the Canadian Nutrient File with USDA data. This finding suggests that match rates partially reflect dietary cultural proximity to the United States rather than database completeness per se. The 45.2% of unmatched USDA entries warrants further investigation. The present study represents a first trial using USDA FDC as a common axis for multi-country food matching. An alternative approach for constructing a truly global food composition reference would be to aggregate all national FCDs into a union dataset, cluster food items by nutrient profile similarity, merge entries that are nutritionally equivalent across countries while retaining their original food names and country of origin, and treat genuinely distinct items as separate entries. Such a union-based approach would be more inclusive than a USDA-anchored one, but would require resolving conflicts in measurement methodology and analytical standards across databases (Sichert-Hellert et al., 2011). If international harmonization of food composition data is to be pursued through FAO/INFOODS frameworks (FAO/INFOODS, 2012) or similar standards, these findings suggest that USDA FDC alone provides an insufficient foundation, particularly for Asian and non-English-speaking dietary contexts.

Historically, efforts such as the EuroFIR project required nearly a decade of expert collaboration (2005–2013) to harmonize food composition data across European countries. The present study demonstrates that LLM-based matching can accomplish in a single overnight computation what would require years of expert manual curation under traditional approaches. As food composition databases continue to proliferate globally, LLM-assisted harmonization can make the construction of a truly universal food composition reference far more tractable than previously possible.

## 5. Conclusion

These findings suggest that while USDA FDC represents a practical foundation for a globally comprehensive food composition database given its breadth, systematic incorporation of country-specific foods and classification schemes will be necessary to achieve true international interoperability.

## Data Availability

The USDA FoodData Central data are available at https://fdc.nal.usda.gov/. National food composition databases used in this study are publicly available from the respective government agencies: Japan (https://www.mext.go.jp/), United Kingdom (https://www.gov.uk/), Australia (https://www.foodstandards.gov.au/), New Zealand (https://www.plantandfood.com/), Canada (https://www.canada.ca/), and South Korea (https://www.foodsafetykorea.go.kr/). Analysis code will be made available on GitHub upon acceptance.

https://www.mext.go.jp/

https://www.gov.uk/

https://www.foodstandards.gov.au/

https://www.canada.ca/

https://www.foodsafetykorea.go.kr/

## Acknowledgements

The authors thank the agencies responsible for developing and maintaining the national food composition databases used in this study. LLM-based matching was performed using the Anthropic Claude API.

## Data Availability

Matching pipeline scripts and prompt templates will be made publicly available at https://github.com/shnkgw-rincom/jbfd-correspondence-table upon finalization.

## Declaration of Generative AI and AI-Assisted Technologies in the Manuscript Preparation Process

During the preparation of this work the authors used Claude (Anthropic) for LLM-based food item matching and manuscript drafting assistance. After using this tool, the authors reviewed and edited the content as needed and take full responsibility for the content of the published article.

